# Modeling COVID-19 Transmission in Africa: Country-wise Projections of Total and Severe Infections Under Different Lockdown Scenarios

**DOI:** 10.1101/2020.09.04.20188102

**Authors:** Isabel Frost, Jessica Craig, Gilbert Osena, Stephanie Hauck, Erta Kalanxhi, Emily Schueller, Oliver Gatalo, Yupeng Yang, Katie Tseng, Gary Lin, Eili Klein

## Abstract

**Objectives:** As of August 24^th^ 2020, there have been 1,084,904 confirmed cases of SARS-CoV-2 and 24,683 deaths across the African continent. Despite relatively lower numbers of cases initially, many African countries are now experiencing an exponential increase in case numbers. Estimates of the progression of disease and potential impact of different interventions are needed to inform policy making decisions. Herein, we model the possible trajectory of SARS-CoV-2 in 52 African countries under different intervention scenarios.

**Design:** We developed a compartmental model of SARS-CoV-2 transmission to estimate the COVID-19 case burden for all African countries while considering four scenarios: no intervention, moderate lockdown, hard lockdown, and hard lockdown with continued restrictions once lockdown is lifted. We further analyzed the potential impact of COVID-19 on vulnerable populations affected by HIV/AIDS and TB.

**Results:** In the absence of an intervention, the most populous countries had the highest peaks in active projected number of infections with Nigeria having an estimated 645,081 severe infections. The scenario with a hard lockdown and continued post-lockdown interventions to reduce transmission was the most efficacious strategy for delaying the time to the peak and reducing the number of cases. In South Africa projected peak severe infections increase from 162,977 to 203,261, when vulnerable populations with HIV/AIDS and TB are included in the analysis.

**Conclusion:** The COVID-19 pandemic is rapidly spreading across the African continent. Estimates of the potential impact of interventions and burden of disease are essential for policy makers to make evidence-based decisions on the distribution of limited resources and to balance the economic costs of interventions with the potential for saving lives.

**ARTICLE SUMMARY:** *Strengths and limitations of this study:*

- Though the rapid spread of SARS-CoV-2 through China, Europe and the United States has been well-studied, leading to a detailed understanding of its biology and epidemiology, the population and resources for combatting the spread of the disease in Africa greatly differ to those areas and require models specific to this context.
- Few models that provide estimates for policymakers, donors, and aid organizations focused on Africa to plan an effective response to the pandemic threat that optimizes the use of limited resources.
- This is a compartmental model and as such has inherent weaknesses; including the possible overestimation of the number of infections as it is assumed people are well mixed, despite many social, physical and geographical barriers to mixing within countries.
- Peaks in transmission are likely to occur at different times in different regions, with multiple epicenters.
- This model is not stochastic and case data are modeled from the first twenty or more cases, each behaving as an average case; in reality, there are no average cases; some individuals are likely to have many contacts, causing multiple infections, and others to have very few.

## INTRODUCTION

On March 11, 2020, the World Health Organization declared the novel severe acute respiratory syndrome coronavirus 2 (SARS-CoV-2) outbreak a pandemic. As of 24^th^ August 2020, there have been over 23,472,067 cumulative confirmed cases of coronavirus disease (COVID-19) and over 809,747 deaths reported globally.[1] The first confirmed COVID-19 case in Africa occurred in Egypt on February 14, 2020. To-date, African countries have reported lower disease incidence than most other countries, with 1,084,904 confirmed cases and 24,683 deaths as of 24^th^ August 2020 across the continent. However, infectious disease surveillance and reporting infrastructure remain highly underdeveloped, and COVID-19 testing is limited given the shortage of human resources and appropriate laboratory and surveillance facilities across the continent.[2]

Although uncertainties underlying SARS-CoV-2 disease transmission and severity persist, ongoing analysis of available data suggests that old age and underlying health conditions play a critical role in the severity of disease prognosis.[3,4] Approximately 75% of the African population is less than 35 years of age, and African countries may benefit from a largely young population.[5] However, these populations may also be at particular risk for high morbidity and mortality from COVID-19 given the high prevalence of immunocompromised individuals. In 2016, 417,000 people died from tuberculosis in the African region, where 25% of the world’s TB deaths occur.[6] Other prevalent co-morbidities include HIV, for which many patients are receiving anti-retroviral therapy, and malnutrition, in addition to other communicable and noncommunicable diseases.[7–9]

To mitigate the spread of COVID-19, the majority of African countries reduced or banned international travel and instituted curfews, lockdowns, and other social distancing interventions beginning in March and April 2020.[10] Several studies have demonstrated the effectiveness of social distancing and other quarantine measures in tandem with a rapid scale up of SARS-CoV-2 testing.[11] By one estimate, the number of people likely to be infected with the virus after encountering an infected individual declined by 55 percent after one week of shutdown in Wuhan, China.[12] However, as the majority of these restrictions were lifted in June 2020, the transmission rate is expected to increase, especially in highly populated areas where social distancing is not feasible.

In this analysis, we combine best available estimates of the parameters that govern SARS-CoV-2 transmission dynamics with country-specific population data to estimate potential COVID-19 case burdens under four scenarios: baseline (assuming disease transmission is not mitigated by appropriate interventions), moderate lockdown, hard lockdown, and hard lockdown with continued social distancing. Given the current lack of data and understanding of how COVID-19 will impact the continent, these estimates are generated to inform national and continental COVID-19 preparation and response efforts on the basis of how this disease has spread elsewhere.

## METHODS

### Data

The number of confirmed COVID-19 cases and the date of the first twenty or more cases were obtained for each country using data aggregated by the Johns Hopkins Centers for Systems Science and Engineering as of 25^th^ June 2020.[1] Lockdown start and end dates, where applicable, were compiled from a variety of sources including news media reports and government statements (Appendix 1 and Table 2). Although data are as current as possible, many government policies are under continuous review and lockdown dates may become rapidly outdated. Where countries do not have dates for the lifting of lockdown, a duration of 60 days has been assumed in line with the recommendations of many governments. Country population estimates were obtained from the World Bank.[13] Projections were simulated across all African nations and territories (excluding Comoros, Mayote, Reunion, Lesotho, and St. Helena which were omitted for lack of sufficient case data).

For all included countries, we used case data for 200 days after the first 20 or more confirmed cases were recorded. Only the Seychelles has not yet reached this 20-case threshold, where confirmed cases remain at 11 since April 6, 2020. For this case the highest number and the date this figure was first recorded were used to initiate the model. Model start and end dates for each country are provided in Table 2.

To incorporate the varying age structures of different countries into the model, parameters were weighted by the proportion of the population in the 0-64, 65-79, and 80 and above age brackets in each country to form a unique set of parameters for each country (Appendix 2). Demographic data were obtained from the World Bank for 2018,[13] with the exception of Eritrea, for which population data came from IndexMundi.[14] Many African countries have high rates of TB and HIV/AIDS which are likely to make their populations more vulnerable to severe infection. To further consider the impact of this on our analysis we reweighted the parameters according to the proportion of the younger population with HIV and/or TB. Those over age 80, and 65-79 were modeled as before, however the under 64 cohort was split into healthy individuals and those affected by TB and/or HIV/AIDS. For this group we employed the parameter set previously used for the 0-64-year-old cohort, with the exception of progression to severe disease, which was doubled to 0·404 (upper bound tripled to 0·606 and lower bound of 0·202), for populations with HIV/AIDS and/or TB, based on estimates for mortality from South Africa.[15,16] Algeria, Mauritius, Eritrea, and Seychelles were excluded from this second analysis due to lack of data.

### Model Structure

#### Epidemiological equations and parameters

The model follows a modified SEIR structure (Figure 1) with seven unique compartments to describe the epidemiology of SARS-CoV-2. This is adapted from the model structure previously described in Lin *et al*. 2020.[17] Susceptible individuals, *S*, are those in the population that can become infected with the virus. They become exposed, *E*, to SARS-CoV-2 by encountering infected individuals in the population at rate β_*1*_ for symptomatic individuals or β_*2*_for symptomatic individuals. We assume that individuals that are asymptomatic or mildly symptomatic have a lower transmission rate, β_*1*_ for asymptomatic individuals or β_*2*_ than more symptomatic individuals,β_*2*_.[17] Exposed individuals incubate the virus at rate μ(calculated as the inverse of the incubation period). A proportion of these individuals,θ, become symptomatically infected while the rest become contagious with mild or no symptoms, *C*. Of the symptomatically infected individuals, a proportion, *h*, have severe symptoms, to the extent that they will require hospitalization if available, *I_S_*, and the rest have moderate or non-severe symptoms, *I_N_*. Asymptomatic or mildly symptomatic, moderately symptomatic, and severely symptomatic individuals recover (or otherwise become non-contagious) at rates *γ_1_,γ_2_* and *γ_3_* respectively. It is assumed that recovered individuals are immune from becoming re-infected during the time period of the study. Severely infected individuals may also die, *D*, at rate δ.

**Figure 1.**
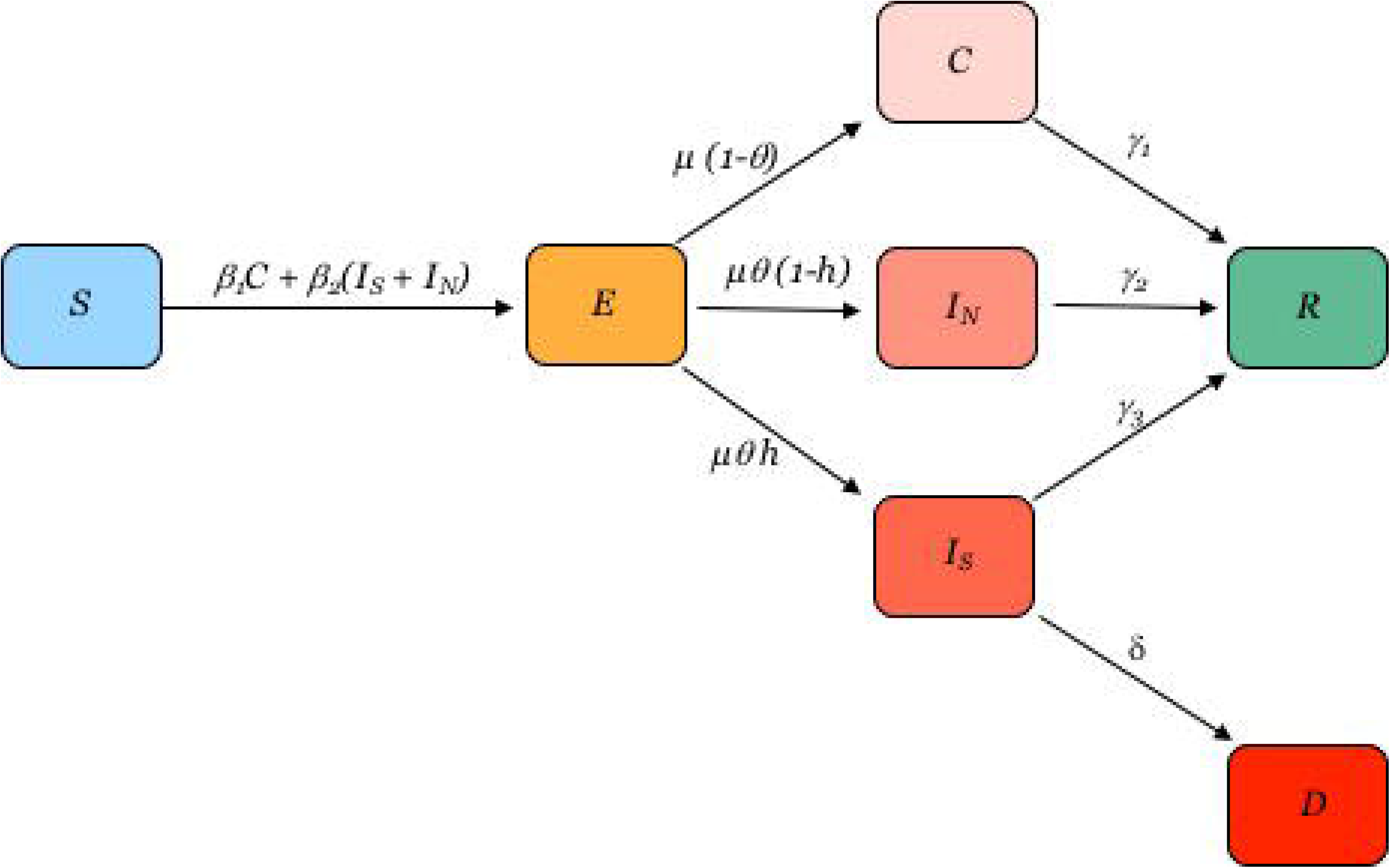
Modified SEIR model structure. Susceptible individuals, *S*, become exposed, *E*, to SARS-CoV-2. A proportion of these individuals become symptomatically infected with severe symptoms, *I_S_*, or non-severe, symptoms, *I_N_*, while the rest become contagious with mild or no symptoms, *C*. Asymptomatic or mildly symptomatic, moderately symptomatic and severely symptomatic individuals recover, *R*, and severely infected individuals may also die, *D*.

The model is described by the following set of differential equations:

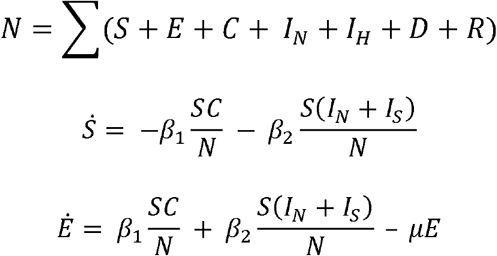

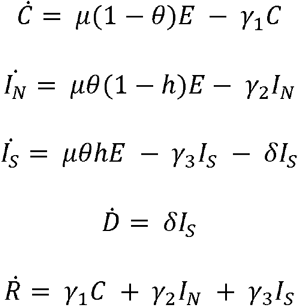

### Estimation of Epidemiological Parameters

Parameters are estimated from the literature and the sources for this are outlined in Table 1 and the supplementary materials.

To assess the uncertainty of the parameter ranges on model estimations, we used Latin Hypercube Sampling (LHS), a stratified sampling technique that efficiently analyzes large numbers of input parameters by treating each parameter as a separate random variable. Stochastic sampling of the parameters with LHS was based on an estimation of parameter ranges obtained from the literature (Table 1). From the parameter sampling, we were able to calculate the 95% confidence interval (CI) for all compartment values over the temporal domain.

**Table 1.**
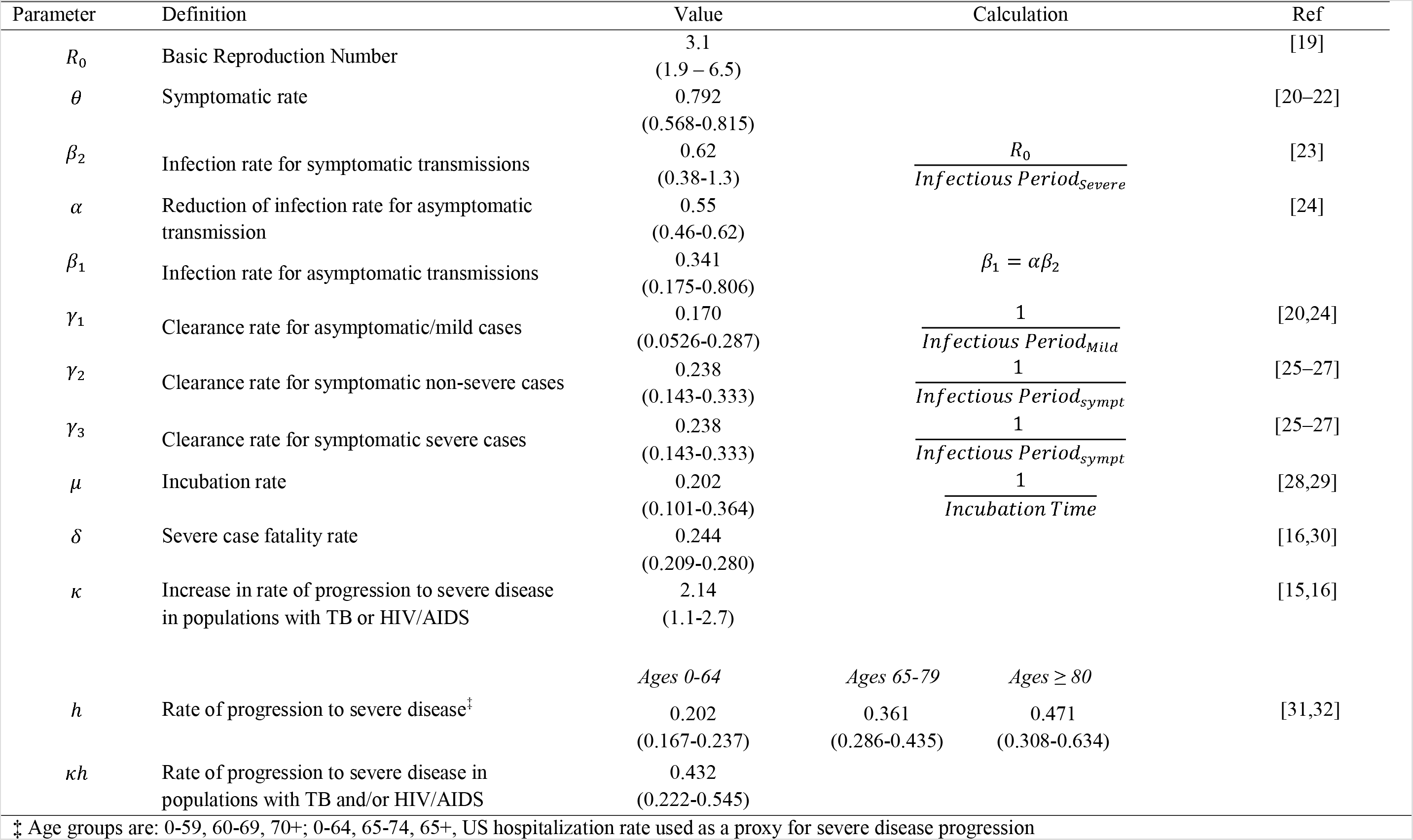
Parameters estimation from other literature where the means and corresponding credible intervals are shown in the parenthesis [19] [20–22] [23] [24] [20,24] [25℃27] [25℃27] [28,29] [16,30] [15,16] [31,32]

| Parameter | Definition | Value | Calculation |  |  | Ref |
| --- | --- | --- | --- | --- | --- | --- |
| $R_0$ | Basic Reproduction Number | 3.1<br>(1.9 – 6.5) | | | | [19] |
| $\theta$ | Symptomatic rate | 0.792<br>(0.568-0.815) | | | | [20–22] |
| $\beta_2$ | Infection rate for symptomatic transmissions | 0.62<br>(0.38-1.3) | $\frac{R_0}{Infectious\ Period_{severe}}$ | | | [23] |
| $\alpha$ | Reduction of infection rate for asymptomatic transmission | 0.55<br>(0.46-0.62) | | | | [24] |
| $\beta_1$ | Infection rate for asymptomatic transmissions | 0.341<br>(0.175-0.806) | $\beta_1 = \alpha\beta_2$ | | | |
| $\gamma_1$ | Clearance rate for asymptomatic/mild cases | 0.170<br>(0.0526-0.287) | $\frac{1}{Infectious\ Period_{Mild}}$ | | | [20,24] |
| $\gamma_2$ | Clearance rate for symptomatic non-severe cases | 0.238<br>(0.143-0.333) | $\frac{1}{Infectious\ Period_{sympt}}$ | | | [25–27] |
| $\gamma_3$ | Clearance rate for symptomatic severe cases | 0.238<br>(0.143-0.333) | $\frac{1}{Infectious\ Period_{sympt}}$ | | | [25–27] |
| $\mu$ | Incubation rate | 0.202<br>(0.101-0.364) | $\frac{1}{Incubation\ Time}$ | | | [28,29] |
| $\delta$ | Severe case fatality rate | 0.244<br>(0.209-0.280) | | | | [16,30] |
| $\kappa$ | Increase in rate of progression to severe disease in populations with TB or HIV/AIDS | 2.14<br>(1.1-2.7) | | | | [15,16] |
|  |  | *Ages 0-64* | *Ages 65-79* | *Ages $\geq 80$* |  |  |
| $h$ | Rate of progression to severe disease<sup>‡</sup> | 0.202<br>(0.167-0.237) | 0.361<br>(0.286-0.435) | 0.471<br>(0.308-0.634) | | [31,32] |
| $\kappa h$ | Rate of progression to severe disease in populations with TB and/or HIV/AIDS | 0.432<br>(0.222-0.545) | | | | |

‡ Age groups are: 0-59, 60-69, 70+; 0-64, 65-74, 65+, US hospitalization rate used as a proxy for severe disease progression

### Scenarios and Assumptions

We provide case projections for the following four scenarios:

1. **Baseline**: Disease continues to spread with no curfew, lockdown, social distancing, or other intervention(s) and with no change in transmission rate.
2. **Moderate Lockdown**: Disease transmission is reduced by 25% during the lockdown period, then transmission resumes at 90% of the pre-lockdown value due to sustained changes in behavior.
3. **Hard Lockdown**: Disease transmission falls 44% during the lockdown period, then transmission resumes at 90% of pre-lockdown levels.
4. **Hard Lockdown and Continued Social Distancing/Isolating Cases**: Disease transmission is reduced by 44% during the lockdown period then, through social distancing regulations and isolation of symptomatic individuals, resumes at 75% of pre-lockdown levels.

For each scenario, we estimate the total number of infections that are asymptomatic or mildly symptomatic, moderately symptomatic, and severely symptomatic cases. The rate of severely symptomatic cases is based on the rate of hospitalization in other parts of the world, although access to hospital care is likely to differ greatly between different parts of Africa.[18]

### Patient and Public Involvement Statement

It was not appropriate or possible to involve patients or the public in the design, or conduct, or reporting, or dissemination plans of our research.

## RESULTS

Under baseline conditions, the most populous countries stand to bear the greatest disease burden with Nigeria having an estimated peak case load of 645,081 severe infections (0·31 percent of total country population) and 9,359,221 total infections (4·54% of total country population), followed by Ethiopia, with an estimated peak case load of 335,024 (0·29%) severe infections and 4,978,734 (4·33%) total infections (Figure 2; Table 3). Smaller countries have a lower case load; Cabo Verde is projected to have a peak case load of 2,244 severe infections (0·40% of total country population) and 32,811 (5·90%) total infections, Sao Tome and Principe are estimated to experience peaks of 14,012 (6·40%) total infections and 1,048 (0·48%) severe infections (Figure S1). However, the baseline scenario does not reflect the current situation in any country as all countries have instituted some form of social distancing policies.

Moderate lockdowns (assumed to lower transmission by 25% during lockdown), reduced estimated peak severe infections by 10% in Senegal, to 37%, in Ethiopia and Egypt. However, South Africa the peak of the severe case load showed a 1% increase of 1,929, given a moderate lockdown of 35 days (Figures S1 and S6). Longer lockdowns were more effective. In Egypt and Ethiopia, who have the longest planned lockdowns of any of the African countries (137 and 170 days, respectively), the estimated impact was a reduction in total peak cases of 37%, or 130,998 and 123,890 severe cases, respectively (Figure 5). In addition, the estimated peak of infections was shifted by 35 and 27 days, respectively (Table 2). In Cabo Verde, which had the shortest planned lockdown of 17 days, the estimated reduction in severe peak cases was 19% under a moderate lockdown, and the peak date of infections was shifted by 10 days (Figure 5).

**Table 2:** Lockdown and Peak Dates

| Country | Beginning Date of Simulation | Start Date of Lockdown | Duration of Lockdown (Days) | Date of Peak Infections (Baseline) | Date of Peak Infections (Moderate Lockdown) | Date of Peak Infections (Hard Lockdown) | Date of Peak Infections (Hard Lockdown with Social Distancing) |
| --- | --- | --- | --- | --- | --- | --- | --- |
| Algeria | 3/9/20 | 3/23/20 | 82 | 7/2/20 | 7/22/20 | 8/27/20 | 8/14/20 |
| Angola | 4/18/20 | 3/31/20 | 70 | 8/6/20 | 8/29/20 | 9/12/20 | 10/3/20 |
| Benin | 4/5/20 | 3/30/20 | 41 | 7/15/20 | 7/31/20 | 8/15/20 | 9/23/20 |
| Botswana | 4/19/20 | 4/6/20 | 46 | 7/16/20 | 8/18/20 | 8/22/20 | 9/5/20 |
| Burkina Faso | 3/18/20 | 3/21/20 | 54 | 7/1/20 | 7/23/20 | 7/25/20 | 8/6/20 |
| Burundi | 5/17/20 | 3/21/20 | 39 | 9/3/20 | 9/11/20 | 9/23/20 | 10/9/20 |
| Cabo Verde | 4/15/20 | 3/28/20 | 17 | 7/6/20 | 7/16/20 | 7/29/20 | 8/3/20 |
| Cameroon | 3/20/20 | 3/18/20 | 44 | 7/6/20 | 7/21/20 | 8/11/20 | 8/29/20 |
| Central African Republic | 4/28/20 | 3/30/20 | 60 | 8/9/20 | 8/31/20 | 9/7/20 | 9/29/20 |
| Chad | 4/13/20 | 4/2/20 | 43 | 7/28/20 | 8/3/20 | 9/6/20 | 9/26/20 |
| Congo (Brazzaville) | 4/2/20 | 3/31/20 | 61 | 7/12/20 | 7/28/20 | 8/27/20 | 8/26/20 |
| Congo (Kinshasa) | 3/21/20 | 3/18/20 | 78 | 7/9/20 | 8/11/20 | 8/30/20 | 9/9/20 |
| Cote d'Ivoire | 3/23/20 | 3/16/20 | 62 | 7/3/20 | 7/20/20 | 8/16/20 | 8/29/20 |
| Djibouti | 3/31/20 | 3/23/20 | 46 | 6/22/20 | 7/20/20 | 8/4/20 | 8/18/20 |
| Egypt | 3/8/20 | 3/15/20 | 137 | 6/22/20 | 7/27/20 | 10/2/20 | 9/14/20 |
| Equatorial Guinea | 4/12/20 | 3/26/20 | 64 | 7/5/20 | 7/27/20 | 8/13/20 | 8/26/20 |
| Eritrea | 4/2/20 | 3/23/20 | 31 | 7/4/20 | 7/24/20 | 7/30/20 | 8/18/20 |
| Eswatini | 4/18/20 | 3/26/20 | 42 | 7/19/20 | 7/28/20 | 8/24/20 | 8/30/20 |
| Ethiopia | 3/29/20 | 3/22/20 | 170 | 7/22/20 | 8/18/20 | 10/27/20 | 11/22/20 |
| Gabon | 4/2/20 | 3/20/20 | 58 | 7/5/20 | 7/19/20 | 8/5/20 | 8/27/20 |
| Gambia | 5/11/20 | 3/17/20 | 59 | 8/11/20 | 8/29/20 | 9/29/20 | 10/8/20 |
| Ghana | 3/22/20 | 3/17/20 | 34 | 7/11/20 | 7/8/20 | 7/25/20 | 8/8/20 |
| Guinea | 3/30/20 | 3/26/20 | 50 | 7/8/20 | 7/19/20 | 8/15/20 | 8/30/20 |
| Guinea-Bissau | 4/7/20 | 3/17/20 | 55 | 7/13/20 | 7/31/20 | 8/12/20 | 8/19/20 |
| Kenya | 3/24/20 | 3/15/20 | 43 | 7/14/20 | 8/14/20 | 8/27/20 | 9/13/20 |
| Liberia | 4/8/20 | 3/22/20 | 21 | 7/15/20 | 7/19/20 | 8/9/20 | 8/18/20 |
| Libya | 4/7/20 | 4/17/20 | 20 | 7/19/20 | 7/28/20 | 8/6/20 | 8/10/20 |
| Madagascar | 3/26/20 | 3/23/20 | 60 | 7/6/20 | 8/5/20 | 8/15/20 | 8/23/20 |
| Malawi | 4/22/20 | 3/23/20 | 48 | 8/7/20 | 8/13/20 | 9/17/20 | 10/21/20 |
| Mali | 3/30/20 | 3/15/20 | 55 | 7/24/20 | 8/8/20 | 8/26/20 | 9/4/20 |
| Mauritania | 5/14/20 | 3/15/20 | 60 | 8/18/20 | 9/9/20 | 10/3/20 | 10/9/20 |
| Mauritius | 3/22/20 | 3/29/20 | 64 | 6/20/20 | 7/15/20 | 8/1/20 | 8/14/20 |
| Morocco | 3/15/20 | 3/15/20 | 87 | 6/28/20 | 7/26/20 | 8/25/20 | 8/18/20 |
| Mozambique | 4/10/20 | 3/23/20 | 68 | 7/20/20 | 8/17/20 | 9/10/20 | 10/9/20 |
| Namibia | 4/5/20 | 3/17/20 | 76 | 7/8/20 | 8/5/20 | 8/17/20 | 8/18/20 |
| Niger | 3/30/20 | 3/20/20 | 113 | 7/14/20 | 8/23/20 | 9/22/20 | 9/16/20 |
| Nigeria | 3/21/20 | 3/30/20 | 63 | 7/15/20 | 8/4/20 | 9/4/20 | 9/30/20 |
| Rwanda | 3/23/20 | 3/21/20 | 60 | 7/2/20 | 7/15/20 | 8/19/20 | 9/12/20 |
| Sao Tome and Principe | 5/5/20 | 3/17/20 | 60 | 7/22/20 | 8/13/20 | 9/1/20 | 9/8/20 |
| Senegal | 3/15/20 | 3/14/20 | 77 | 6/30/20 | 7/17/20 | 8/12/20 | 8/18/20 |
| Seychelles | 4/6/20 | 3/16/20 | 60 | 6/19/20 | 7/5/20 | 7/21/20 | 8/15/20 |
| Sierra Leone | 4/17/20 | 4/4/20 | 60 | 7/21/20 | 8/8/20 | 8/28/20 | 9/13/20 |
| Somalia | 4/10/20 | 3/18/20 | 60 | 7/17/20 | 8/17/20 | 8/24/20 | 9/15/20 |
| South Africa | 3/13/20 | 3/27/20 | 35 | 6/23/20 | 7/11/20 | 8/11/20 | 8/1/20 |
| South Sudan | 4/28/20 | 3/25/20 | 60 | 8/9/20 | 9/7/20 | 9/9/20 | 9/21/20 |
| Sudan | 4/13/20 | 3/15/20 | 80 | 7/29/20 | 8/22/20 | 9/19/20 | 10/9/20 |
| Tanzania | 4/1/20 | 3/23/20 | 60 | 7/18/20 | 8/3/20 | 9/3/20 | 9/20/20 |
| Togo | 3/24/20 | 3/20/20 | 60 | 7/2/20 | 7/12/20 | 8/30/20 | 8/16/20 |
| Tunisia | 3/16/20 | 3/12/20 | 53 | 6/20/20 | 7/31/20 | 7/27/20 | 8/12/20 |
| Uganda | 3/27/20 | 3/18/20 | 47 | 7/10/20 | 7/30/20 | 8/16/20 | 8/27/20 |
| Zambia | 3/27/20 | 3/17/20 | 38 | 6/30/20 | 7/27/20 | 8/15/20 | 9/3/20 |
| Zimbabwe | 4/15/20 | 3/23/20 | 60 | 7/26/20 | 8/25/20 | 9/8/20 | 9/19/20 |

The effect of hard temporary lockdowns without extended post-lockdown social distancing compared to moderate lockdowns varied by country. The reduction in severe peak infections ranged from 7%, in Rwanda, to 35% in Egypt, compared to baseline values. In some countries a hard lockdown had a lower impact on reducing peak cases than a moderate one. For example, in Ethiopia the expected impact of a moderate lockdown reduced severe cases by 123,890 cases compared to baseline, while a hard lockdown reduced cases by 100,743. However, hard lockdowns delayed the onset of the peak in infections compared to moderate lockdowns by several weeks. For example, in Tanzania, a moderate lockdown of 60 days delayed the peak in severe infections by 16 days compared to a 47-day delay in the case of a hard lockdown.

For all countries, hard lockdowns with continued post-lockdown interventions were the most effective in delaying and reducing peak infections. Delays to the peak in infections ranged from 22 days in Libya to 123 days in Ethiopia (Table 2). This reflects the different lengths of lockdowns in these countries which lasted 20, and 170 days respectively. Hard lockdowns with continued interventions also led to the greatest reductions in the peak in estimated total infections to the greatest extent in these model projections, from 36% in Ghana and South Africa to 58% in Namibia. In Kenya this would reduce the peak case load in severe infections by 100,552 cases with a lockdown of 43 days.

Many African countries have young populations, who have been less likely to show severe symptoms in other countries, however, the high prevalence of HIV/AIDS and TB in these populations potentially renders them more vulnerable to COVID-19.[15] The highest proportional burden HIV countries in Africa are Eswatini, Botswana and South Africa and the highest proportional burden tuberculosis countries are Burundi and Central African Republic. Assuming individuals with TB and HIV/AIDS, once infected, are more likely to progress to severe disease increased the peak number of infections significantly (Figure 2). In Eswatini, Botswana, and South Africa the baseline peak number of severe infections increased from 4,529 to 5,279, 9,334 to 10,023, and from 162,977 to 203,261, respectively (Figure S2, S11-S18). In Burundi, the peak number of severe cases increased from 40,417 to 44,058. However, progression to severe disease may, under certain circumstances, lead to less infections, as those with severe disease may be more likely to die, quarantine, or be sick enough that they are not widely transmitting the disease. For example, in Nigeria, severe infections decrease from 645,081 to 591,888 under this scenario.

**Figure 2:**
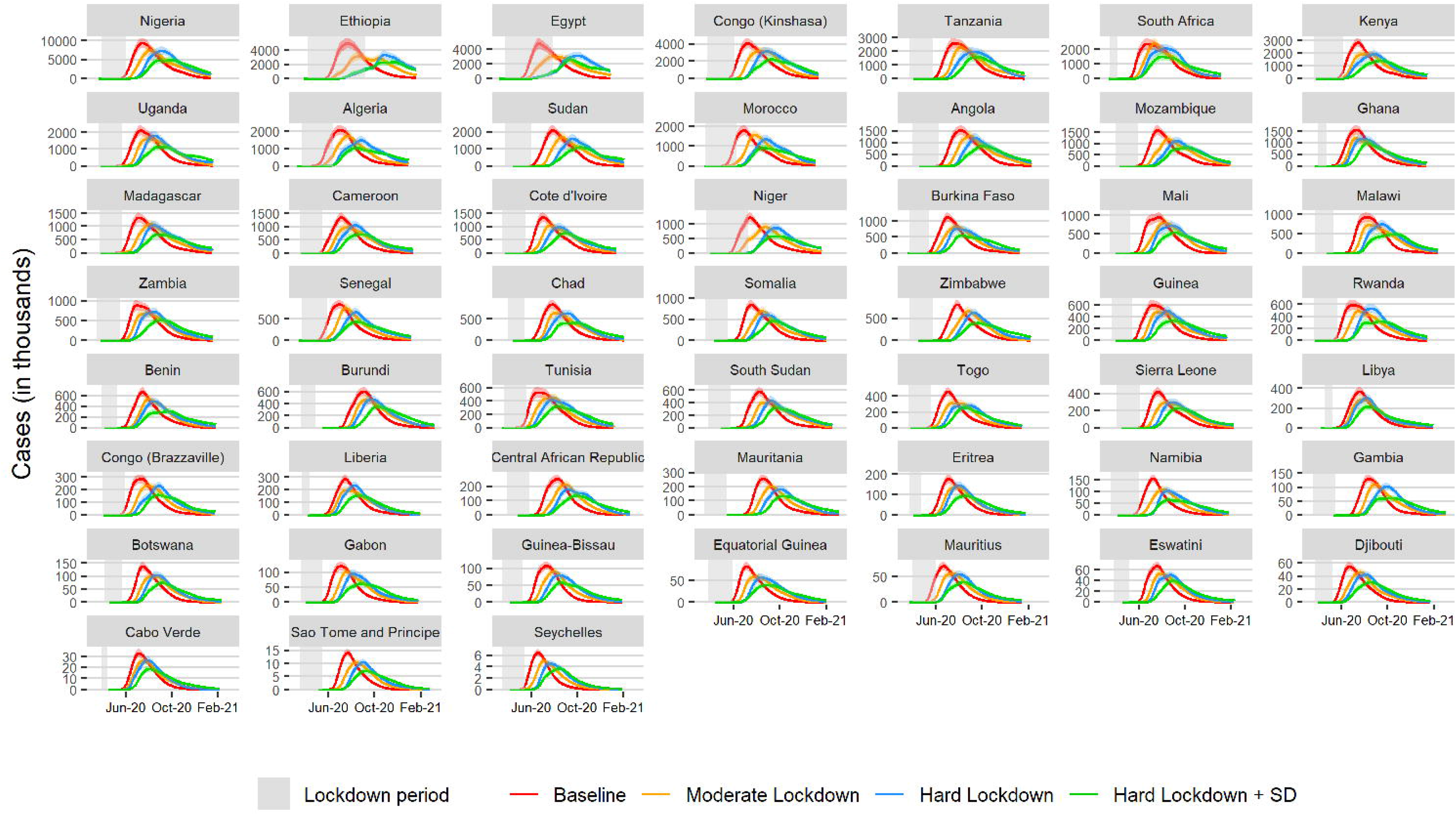
Projected total infections over time when parameters are normalized for the age distribution of the population in each country. Areas shaded in grey denote lockdown duration. SD denotes ‘social distancing.’

**Figure 3:**
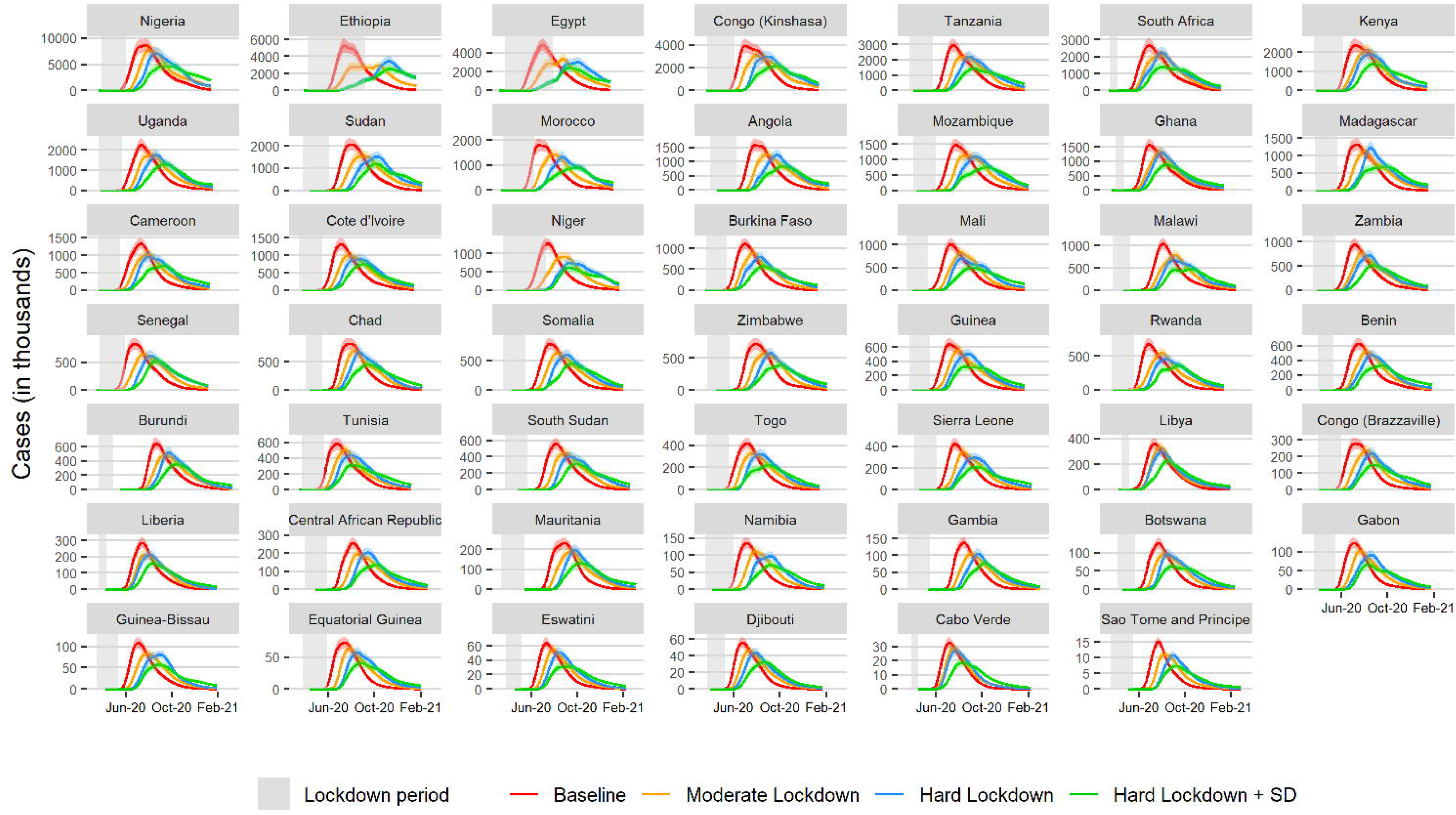
Projected total infections over time when parameters are normalized for the age distribution of the population in each country and the fraction of the under 70-year-old population with HIV/AIDS and or TB. Areas shaded in grey denote lockdown duratio

**Figure 4.**
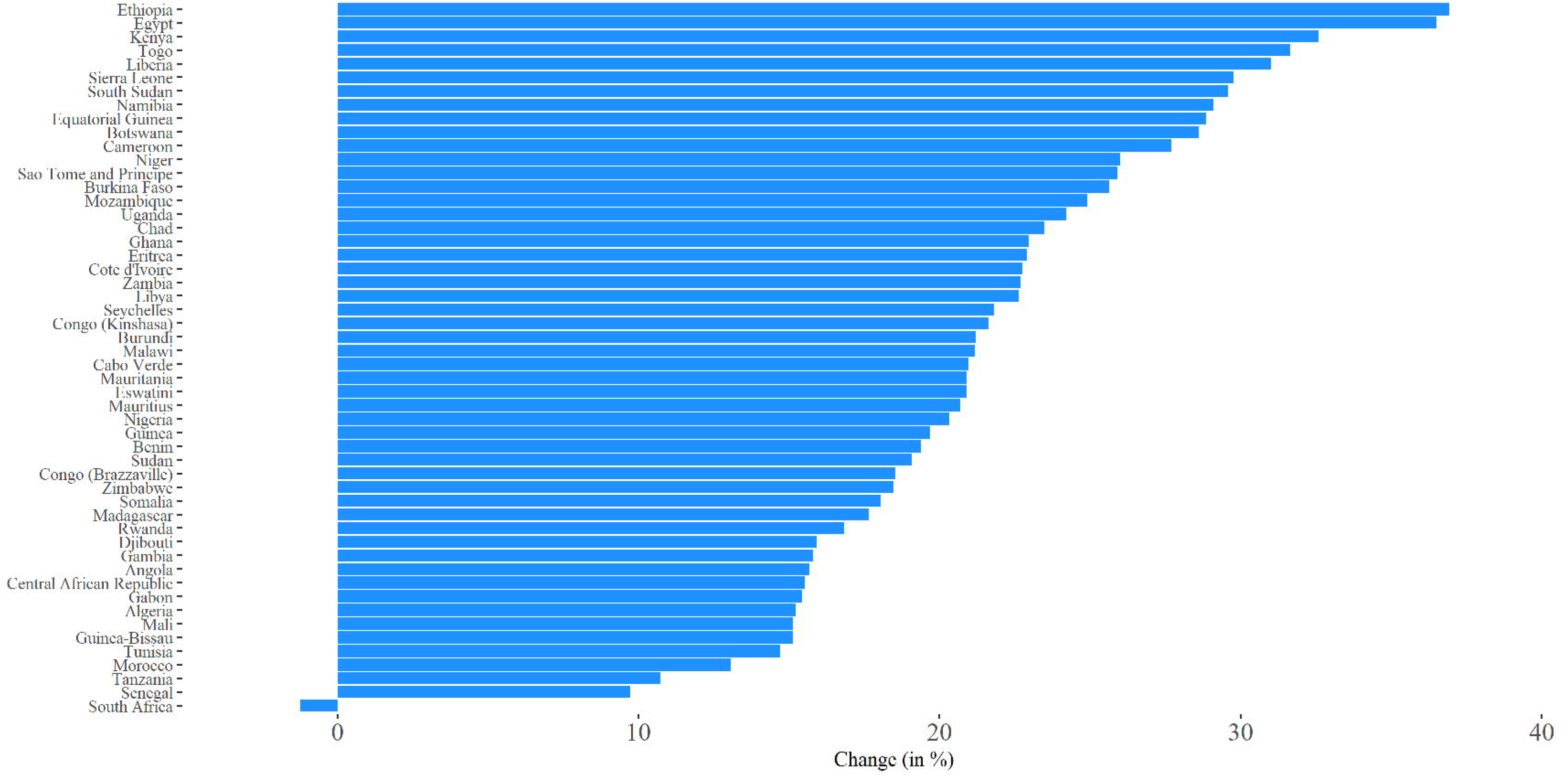
Percent change in peak severe infections under moderate lockdown scenario, parameters adjusted by age only.

**Figure 5.**
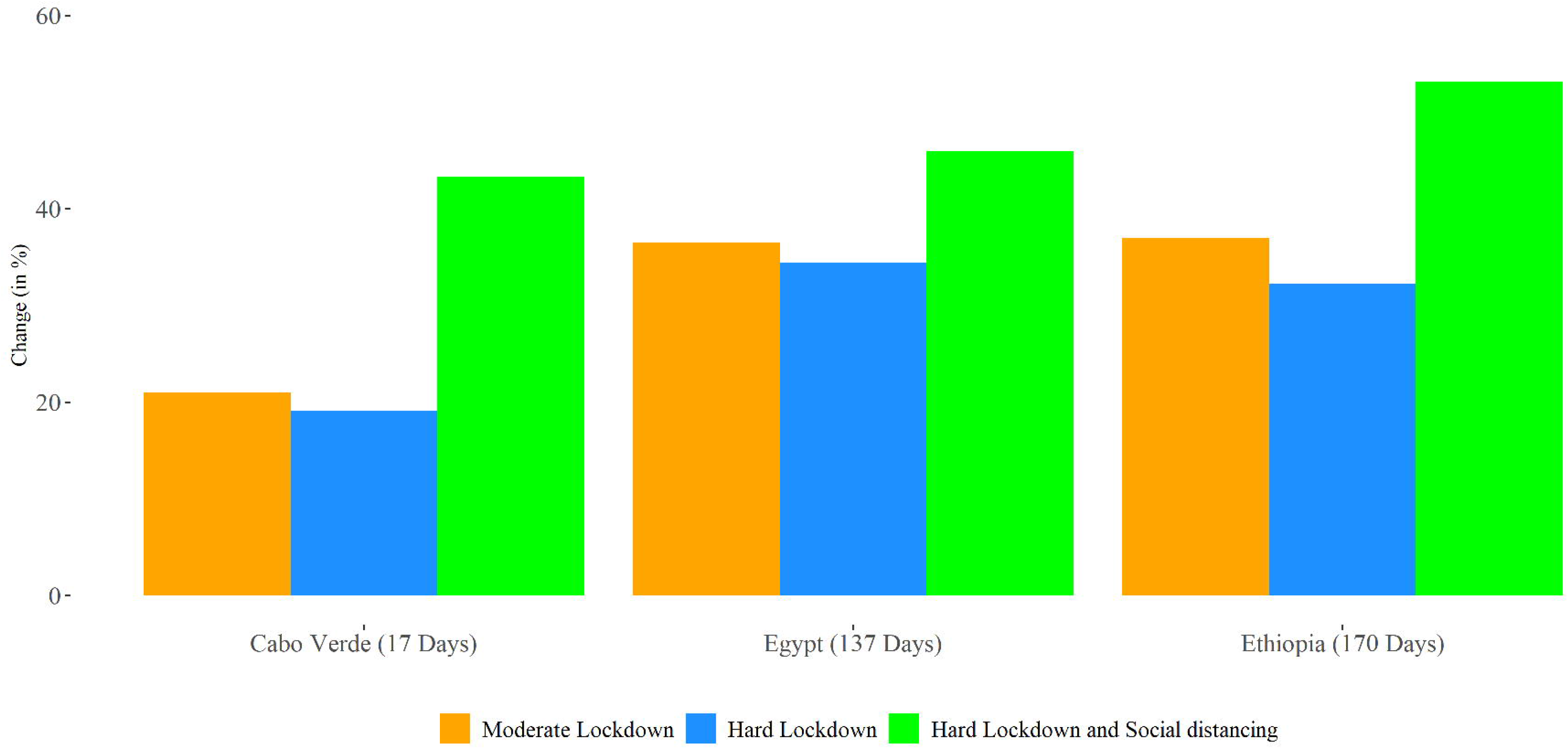
Percent change in peak total infections under interventions, compared to baseline, parameters adjusted by age only. Values in parentheses represent the duration of lockdown in the respective countries. Percent change was calculated relative to a baseline scenario of disease spread with no intervention.

## Conclusion

Most African countries are likely early in the outbreak of SARS-CoV-2, and the initial peak in infections may be several months away in many cases. Policymakers need mathematical models that are attuned to the context in Africa to aid in planning for continued transmission of the virus and to develop interventions that reduce disease transmission. Here we analyzed a model of transmission of SARS-CoV-2 parameterized for low-resource settings. Based on current observed cases of COVID-19 in African countries, we assessed the impact of strict social distancing measures. However, the extent and efficacy of lockdown policies is certain to vary between and within countries. Furthermore, after a lockdown, and in response to a high death toll, citizens are likely to continue to adjust their social behavior.[33] The results from our simulations suggest that national lockdowns will likely slow viral transmission, reducing the peak number of active cases and delaying the time until the peak occurs. This delay can allow governments time to prepare by setting up designated COVID-19 treatment sections in hospitals and additional testing centers in densely populated areas, as well as practical measures within communities such as handwashing stations and soap distribution and information campaigns educating the public in infection prevention behaviors including mask wearing, social distancing and handwashing. Our models suggest that by using this time to prepare, transmission is likely to decrease enough to substantially to reduce the peak in infections, even after lockdowns cease. This may make the consequences of the pandemic more manageable for health and social systems, though many are still likely to become overwhelmed.

The predicted dates of peak cases varied greatly by country, from 1^st^ August in South Africa to 22^nd^ December in Ethiopia in the case of a hard lockdown with continued social distancing (Scenario 3) (Table 6). This is due to many factors including differences in population size, when the virus first arrived in the country, and length and severity of lockdown interventions. Most infections are mild; however, some countries are likely to bear a much higher per capita burden than others, largely due to differences in the duration and efficacy of lockdowns.

This model considers the impact of lockdown on transmission of SARS-CoV-2; however, effective implementation of large-scale mitigation measures thus far implemented in developed countries may not be feasible or sustainable in many low- and middle-income countries (LMICs) in Africa and around the world given various sociocultural, economic, and political challenges.[34,35] Across the African continent, an estimated 40% of people live below the international poverty line making less than $1·90 (in 2011 purchasing parity power) per day, and approximately 85·8% of employment is informal.[36,37] Therefore, lockdowns that restrict movement to and from work will likely not be well enforced. Furthermore, access to hygiene and sanitation facilities is limited; in 2017, only 15% of people across sub-Saharan Africa had access to basic handwashing facilities with soap and water. [38] Social distancing within communities and within households is often not possible given over 55% of Africa’s urban populations live in densely populated slums, higher than the global average of 30%.[39] In addition, there are 6·3 million refugees and 17·7 million internally displaced persons in the African continent, and ongoing humanitarian crises have displaced over 20 million people. These challenges will also abrogate the effectiveness of restrictions on movement and access to care.[40]

COVID-19 presents most severely in the elderly population and those with chronic noncommunicable diseases such as diabetes and hypertension, which affects an estimated 55% of Africans.[41] African populations may benefit from having a younger population and low prevalence of diabetes (3·9%) compared to the global average (9·3%).[42,43] In the model, we modify the transmission and mortality rates according to the age structure of countries, based on evidence that morbidity and mortality is concentrated in older individuals. However emerging reports of COVID-19 cases in the developing world, particularly Brazil, suggest the death toll in the young may be higher than expected.[44] Young populations who go out to work, buy food and look after the family are hard to shield and likely to be highly exposed to the virus. The average size of households with older members is 12·1 in Senegal and 12·6 in the Gambia, the highest in the world,[45] and this may increase the potential for exposure to the virus and hinder isolation of symptomatic cases. High rates of tuberculosis, HIV, malaria, and other infectious diseases may also make young African populations more vulnerable to severe infection with COVID-19. In West and Central Africa 60% of people living with HIV do not receive treatment.[46] The high prevalence of malnutrition, anemia, and exposure to indoor air pollution, often from cooking fires, may also increase vulnerability. Additionally, the poor air quality in many Africa cities, which has been shown to be associated with increased morbidity and mortality from COVID-19,[47,48] may exacerbate issues for the region.[49]

Testing capacity is limited in much of Africa,[50] and confirmed cases may increase faster than predicted in the near future as testing capacity increases and contact tracing continues. In addition, evidence suggests that many cases are asymptomatic and may be missed by testing protocols that only include those with symptoms who have recently travelled to an infected area or their contacts. Furthermore, social stigma and inability to access healthcare may prevent symptomatic individuals from seeking treatment. This may affect the results of the model as it uses the initial numbers of confirmed COVID-19 cases recorded. In addition, results from the model suggest cases may rapidly rise after a hard lockdown if there are no further mitigation measures. Contact tracing and testing are needed to maintain the reduction in cases gained from early lockdowns. Pooled testing can make mass testing more affordable and achievable with limited resources.[51]

There are inherent difficulties in inferring real world results from mass action models such as the one in this study. Our models tend to overestimate the number of infections as they assume people are well mixed, despite many social, physical and geographical barriers to mixing within countries. Peaks in transmission are likely to occur at different times in different regions, as has occurred in the United States and Europe where there have been multiple epicenters. This model is not stochastic and case data are modeled from the first twenty or more cases, each behaving as an average case. In reality, there are no average cases; some individuals are likely to have many contacts, causing multiple infections,[52] and others to have very few. The different contact patterns of different segments of the population were also not included in this model and may have an impact on transmission between vulnerable groups.

The estimates presented here suggest that the burden of severe disease caused by SARS-CoV-2 is likely to be high for the African continent. Projections of disease progression are needed to enable policy makers, governments, aid agencies and other actors to optimize resource allocation and planning decisions. The high prevalence of TB, HIV, and malnutrition and other immunocompromising conditions accompanied by limited testing capacity and access to healthcare in many African countries are likely to make populations particularly vulnerable to this pandemic. Immediate planning and appropriate resource allocation are essential to save lives and mitigate the impact of COVID-19 in Africa.

## Data Availability

Please contact the corresponding author for any queries regarding data availability.

## AUTHOR CONTRIBUTIONS

IF, JC, GO, KT, OG, ES, E. Kalanxhi and SH contributed to the analysis, data collection and figures. E. Klein, GL and YY developed the model and original code with adaptations and additions by IF. IF, JC, and SH contributed to the writing, and all authors provided critical revisions of the manuscript.

## ACKNOWLEDGEMENTS

This work was supported by the Centers for Disease Control and Prevention Modeling in Infectious Disease (MInD) in Healthcare Network.

## DATA SHARING

The data for this study is included in the manuscript and supplementary materials, however please contact the corresponding author for any further data.

